# IN-HOSPITAL MORTALITY AND THE PREDICTIVE ABILITY OF THE MODIFIED EARLY WARNING SCORE IN GHANA

**DOI:** 10.1101/2020.08.05.20169219

**Authors:** Enoch Joseph Abbey, Jennifer Sophie Mammen, Sari Soghoian, Maureen Cadorette, Promise Ariyo

## Abstract

**BACKGROUND:** The modified early warning score (MEWS) is an objective measure of illness severity that promotes early recognition of clinical deterioration in critically ill patients. Its primary use is to; facilitate faster intervention or increase the level of care. Despite, its adoption in some African countries, MEWS is not standard of care in Ghana. We assessed the validity of MEWS as a predictor of mortality, among medically ill in-patients at the Korle-Bu Teaching Hospital (KBTH), Accra – Ghana. We sought to identify the predictive ability of MEWS in detecting clinical deterioration among medical in-patients and its comparability to the routinely measured vital signs.

**METHOD:** This was a retrospective study of medical in-patients, aged ≥13 years and admitted at KBTH from January 2017 – March 2019. Vital signs at 48 hours post-admission were coded using MEWS criteria, to obtain a limited MEWS score (LMEWS) and the level of consciousness imputed to obtain a full MEWS score (MEWS). A predictive model comparing mortality among patients with “significant” MEWS (L/MEWS ≥4) and “non-significant” MEWS (L/MEWS <4) scores was designed using multiple logistic regression. Internally validated for predictive accuracy, using the Receiver Operating Characteristic (ROC) curve.

**RESULTS:** 112 patients were included in the study. The adjusted odds of death comparing patients with a “significant” MEWS to patients with “non-significant” MEWS was 6.33(95% CI 1.96 – 20.48). Similarly, the adjusted odds of death comparing patients with “significant” versus “non-significant” LMEWS was 8.22(95% CI 2.45 – 27.56). The ROC curve for each analysis had a C-static of 0.83 and 0.84 respectively.

**CONCLUSION:** LMEWS is a good predictor of mortality and comparable to MEWS. Adoption of LMEWS can identify medical in-patients at risk of deterioration and death.

## BACKGROUND

Critical illness is a leading cause of morbidity and mortality in Sub-Saharan Africa (SSA) including Ghana. ^[1]^ Low and middle-income countries have a disproportionately higher burden of critical illness with over 90% of global maternal deaths, deaths from trauma, and from infections.^[1-3]^ In Ghana, the critical care burden is high, whereas historically the financial investment has been skewed towards primary health care. Less commitment to critical care means that resources for intensive medical care are limited^[4]^, and their thoughtful and appropriate allocation is important.

One of the main reasons why patients deteriorate and die in hospitals is delayed recognition of illness severity on understaffed in-patient wards. Early warning tools to help identify patients at highest risk of death could help countries like Ghana with resource allocation and clinical decision-making (**Figure 1**).

**Figure 1:**
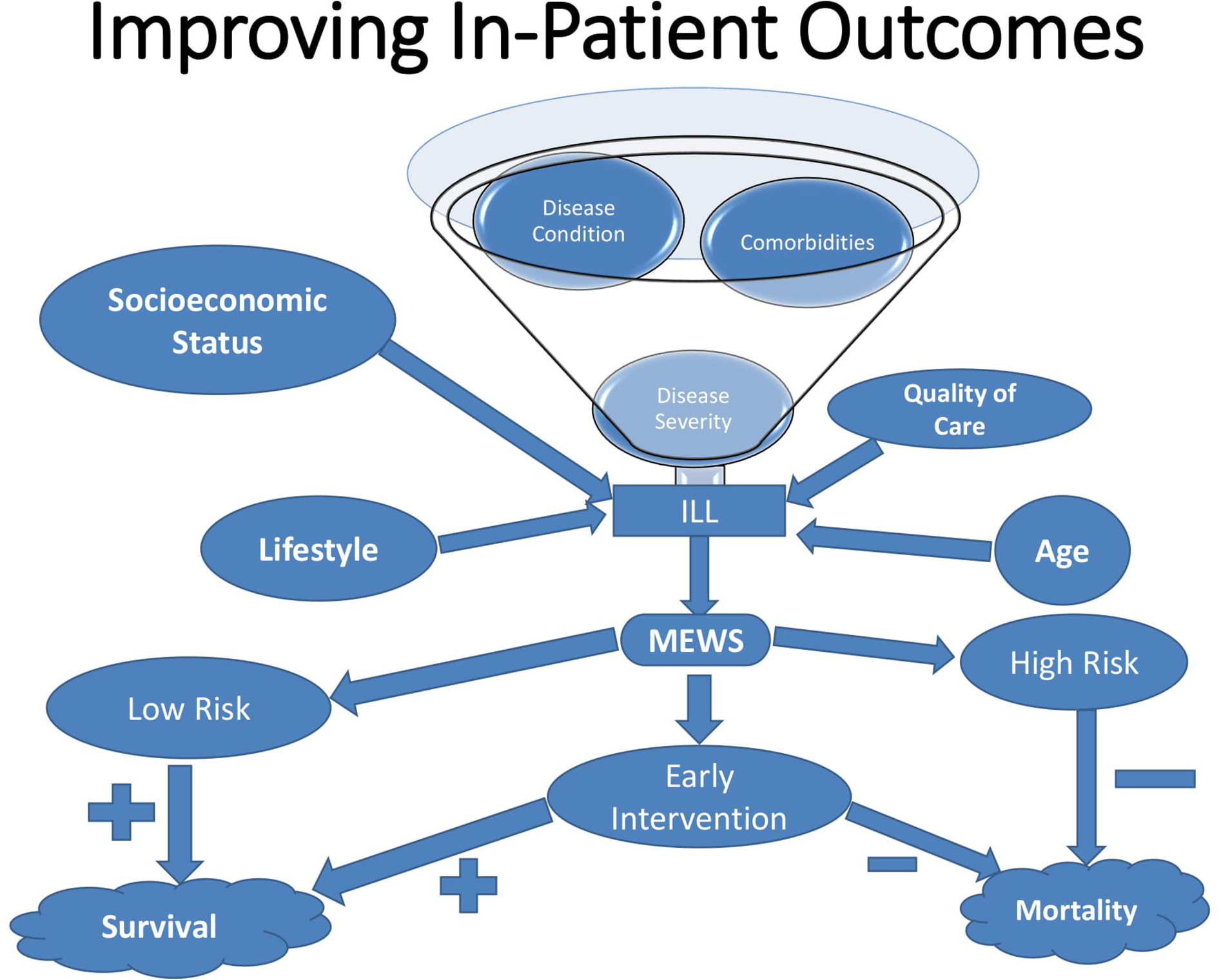
Conceptual Framework showing predictors of in-hospital mortality & role of MEWS

Multiple studies have shown that critical illness and serious adverse events (SAEs) in hospitalized patients are preceded by signs of clinical deterioration in up to 80% of those affected.^[5-8]^ Therefore, changes in physiological parameters can be used to predict adverse events such as shock, cardiac arrest, death, and unplanned intensive care unit (ICU) admissions.^[9]^

The MEWS is a commonly used illness severity score that is calculated by combining five physiologic bedside parameters; systolic blood pressure, heart rate, respiratory rate, temperature and level of consciousness assessed by the AVPU scale (**<u>A</u>**lert, responds to **<u>V</u>**oice, responds to **<u>P</u>**ain & **<u>U</u>**nconscious scale) or RASS score (**<u>R</u>**ichmond **<u>A</u>**gitation-**<u>S</u>**edation **<u>S</u>**cale). These four vital signs and single observation are individually scored and summed to give a combined score between 0 and 14, with higher scores representing increased illness severity.

In a systematic review conducted by Smith et al in 2014, early warning scores, including MEWS had strong predictive ability for death and cardiac arrest within 48 hours in academic urban hospitals for economically advanced countries.^[11]^ Early warning scores have also been shown to provide precise, concise and unambiguous means of identifying and communicating about clinical deterioration to help clinical staff provide special attention and care to patients who need it most (justifiable appropriation of care).^[12]^ As a result, scoring systems such as MEWS have been adopted in most developed countries and some African countries.^[13-15]^

The present study sought to validate the use of MEWS as a clinical decision-making tool to improve early identification of hospitalized medical patients at increased risk for death in Ghana. In addition, since level of consciousness is not routinely recorded in current clinical practice, we aimed to investigate the predictive utility of a limited MEWS calculation based on vital signs alone. Most studies in similar settings find that the level of consciousness is generally high (well-oriented) even when the other aspects of the MEWS score are abnormal.^[2]^ We therefore hypothesized that the physiologic data currently being monitored in Ghana may be sufficient to improve the early detection of critical illness and help guide resource allocation among in-patients in this setting.

## OBJECTIVE

To determine the predictive ability of the MEWS in detecting clinical deterioration in a low-resource setting.

To determine how the MEWS compares with physiologic measures currently monitored in a tertiary facility in Ghana.

## METHODOLOGY

### STUDY DESIGN AND POPULATION

This was a retrospective chart review study of hospitalized medical patients, aged 13 years and above at the Korle-Bu Teaching Hospital (KBTH), in Accra – Ghana. KBTH is the national hospital of Ghana and the leading tertiary care referral center in the country. Medical in-patients hospitalized there for at least 48 hours whose records were still available between the periods of January 2017 and March 2019 were included in the study. Pediatric patients, defined as those aged less than 13 years of age by the Ministry of Health guidelines were not included. Patients with more than one hospital admission in the past month or were admitted for conditions other than medical ones were also excluded.

Demographic data was collected to analyze covariates. Reported vital signs at 48 hours from admission were recoded and scored to generate a limited MEWS (LMEWS), using thresholds as previously described (**Table 1**).^[2]^ To compare the utility of the LMEWS to the full MEWS in the absence of routine observation of consciousness and recording of AVPU scores, we generated a full MEWS (MEWS) score using imputation by randomly assigning 92% of the study sample to a status of “Alert” (AVPU score of 0) and the rest to scores between ‘1 – 3’. These percentages were determined based on the findings of a study in a similar patient population by Subbe et al., 2001.^[2]^

**Table 1:**
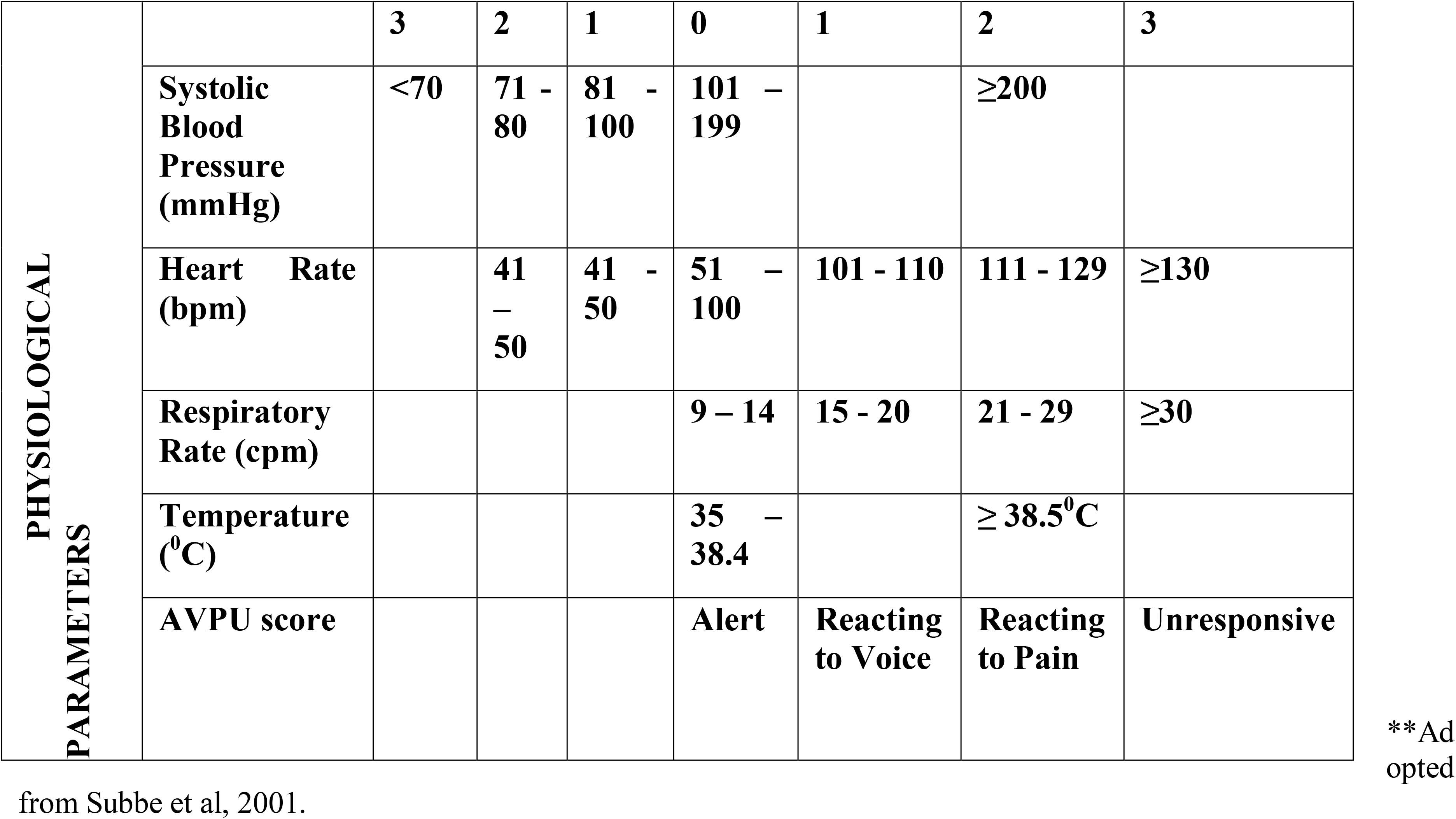
Scoring Scale: Modified Early Warning Score (MEWS) adapted from Subbe et al^2^

The study was based on the conceptual framework depicted in **Figure 1**, which identifies correlational patterns of how different events and experiences may predict mortality in a hospitalized patient. A predictive model was designed using multivariable logistic regression and validated for model accuracy to compare patients with “significant” MEWS to patients with “non-significant” MEWS, where a “significant” MEWS is defined as any score ≥4, and a “non-significant” MEWS is defined as a score <4 based on the absence of the AVPU.^[3, 16, 17]^ This cut-off did not vary for the limited versus imputed scores, because for most individuals the level of consciousness is normal and therefore contributes 0 points to the total MEWS score.

Ethical approval was obtained from the Institutional Review Board (IRB) of Johns Hopkins University, the Scientific and Technical Committee and the IRB of the Korle-Bu Teaching Hospital.

### STATISTICAL ANALYSIS

Data was analyzed using STATA^®^ (Statistical Data Analysis Package version 15.1 IC, College Station, TX – USA). Test for differences between survival to discharge and in-hospital mortality was conducted using a two-sample t-test for continuous physiological parameters, while Chi-square (□^2^) for categorical variables was summarized as frequencies and proportions. Univariable log-binomial regression analysis was used to estimate unadjusted risk ratios between each covariate and mortality. Multivariable Poisson regression with robust variance was used due to the failure of convergence of the log-binomial regression model. Logistic regression analysis (Odds ratio) was used to identify an appropriate predictive model. A p-value of < 0.05 was considered statistically significant. The accuracy of the prediction model was determined using ROC (receiver operator characteristic) curve and C-statistic. Adjustment was made for the following potential confounders; age, sex, duration of admission, admission to the ICU, the presence or absence of other comorbidities and the organ system involved in the disease process.

## RESULTS

The study sample comprised 112 patients admitted for medical reasons during the study period. Of these 62% (69/112) were males with a mean age of 47 (SD ± 17.5) years and 38% were females with a mean age of 52 (SD ± 20) years (**Table 1**). Overall mortality was 41.1%, and increased with age. Every year increase in age was associated with a 3% increase in mortality rate after adjusting for MEWS score [IRR=1 .03 95% CI 1.02 – 1.04]. For patients who survived, the most common admission diagnoses were genitourinary system abnormalities (26.2%), whereas neurologic conditions were most common among patients who died (39%). The longest length of in-hospital stay was 32 days, with an average of 8 days.

The mean systolic blood pressure for patients measured at 48 hours post-admission was 125 mmHg (SD ± 2.9), average pulse rate for patients was 91 mmHg (SD ± 2) mean axillary temperature was 36.9°C (SD ± 0.1) and the average respiratory rate 24 cpm (SD ± 4.7). Only temperature and respiratory rate were individually associated with mortality (**Table 2**). Physiological parameters measured at 48 hours produced an average LMEWS of 3 (range 0-11). Imputation of randomly assigned AVPU values increased mean scores by 8% overall, producing an average MEWS of 3 (range of 0-14).

**Table 2:** Baseline characteristics of cohort

| **Characteristic** | **Survival to Discharge**<br>**(n=66)** | **Death in Hospital (n=46)** | **P-value** |
| --- | --- | --- | --- |
| **Sex (Male)** | **45(68.2)** | **24(52.2)** | **0.087** |
| **Age (years)** |  |  | **<0.001** |
| **25 to 64** | **46 (69.7)** | **27 (58.7)** |  |
| **>=65** | **7 (10.6)** | **18 (39.1)** |  |
| **Disease type by System**<br>**Involved;** |  |  | **0.008** |
| **Cardio-Pulmonary** | **15 (23.1)** | **13 (28.3)** |  |
| **Neuro-Endocrine** | **11 (16.9)** | **18 (39.1)** |  |
| **Hema-Oncology** | **11 (16.9)** | **1 (2.2)** |  |
| **Physiological Parameter at**<br>**48hrs, Mean (SD)** | **Survival to Discharge** | **Mortality in Hospital** | **P-value** |
| **Systolic Blood Pressure**<br>**(mmHg)** | **127.8 (29.4)** | **120.7 (32.1)** | **0.23** |
| **Pulse Rate (bpm)** | **89 (17.6)** | **94 (18.1)** | **0.17** |
| **Axillary Temperature (°C)** | **36.7 (0.7)** | **37.3 (1.2)** | **0.002** |
| **Respiratory Rate (cpm)** | **23 (4.7)** | **25 (6.9)** | **0.0319** |
| **Avg. length of admission** | **7 (6.3)** | **8 (7)** | **0.6** |

\*\* Results are mean (SD) or n (%) P-values obtained via t-test and Chi square ( $\chi^2$ )

**Table 3:** Multivariable Logistic regression of death using Full MEWS & Limited MEWS

| **COVARIATE** | **FULL MEWS - ODDS**<br>**RATIO (95% CI)** | **LIMITED MEWS - ODDS**<br>**RATIO (95% CI)** |
| --- | --- | --- |
| **Age (years)** | 1.08 (1.04 – 1.12) | 1.08 (1.04 – 1.12) |
| **Sex (male)** | 0.44 (0.16 – 1.23) | 0.40 (0.14 – 1.13) |
| **MEWS (Significant)** | 6.33 (1.96 – 20.49) | 8.22 (2.45 – 27.56) |
| **Duration of admission**<br>**(days)** | 0.99 (0.93 – 1.07) | 1.01 (0.94 – 1.08) |
| **Diseased Organ System** | 0.59 (0.31 – 1.13) | 0.59 (0.31 - 1.12) |

“Significant” MEWS was associated with a relative risk of 2.01 (95% CI 1.33 – 3.04) for death on univariable analysis, while a “significant” LMEWS had a relative risk of 2.19 (95% CI 1.46 – 3.30) on univariable analysis.

The death rate calculated by Poisson regression after adjusting for age alone was 2.02 (95% CI 1.40 – 2.91) times higher in patients with a “significant” MEWS compared to those with a “non-significant” MEWS. The death rate for a “significant” MEWS score using LMEWS is 2.13 (95% CI 1.48 – 3.07) times that of “non-significant” MEWS after adjusting for age.

In the multivariable predictive model adjusting for age, sex, duration of admission, admission to the ICU, organ system involved and comorbidities, the odds of death among patients with a ‘significant” MEWS was 6.33 (95% CI 1.96 – 20.50) times that of patients with a non-significant MEWS. The death rate among patients with a “significant” LMEWS was 8.2 (95% CI 2.5 – 27.6) times that of patients with a “non-significant” LMEWS in the multivariable analysis. The best multivariable regression model was selected based on the Akaike Information Criteria (AIC), with a value of 116.4. The odds of death for every year increase in age was 8%, [OR=1.08, 95% CI 1.04 – 1.12]. Other covariates were not statistically significant.

Both LMEWS and MEWS were found to have good discrimination based on the ROC curves, with a C-statistic of 0.838 and 0.833 respectively (**Graphs 1** and **2**) using a cut-off of ≥4. Sensitivity analyses using a “significant” MEWS or LMEWS cut-off value of score ≥ 5 yielded a multivariable odds ratio of 12.4 (95% CI 2.5 – 61.2) and 15.1 (95% CI 2.5 – 91.8) respectively. The ROC curves for LMEWS and MEWS respectively was found to be 0.840 and 0.838 apiece as captured in **Graphs 3** and **4**, when a cut-off of ≥5 was adopted. The Hosmer-Lemeshow goodness-of-fit test yielded Pearson’s chi-square values of 0.254 and 0.163 for LMEWS and MEWS respectively. The Hosmer-Lemeshow test to assess goodness-of-fit yields p-values of 0.766 versus 0.514 for LMEWS and MEWS when a cut-off of ≥5 was used.

## DISCUSSION

MEWS has been validated in several settings as a robust predictor of both clinical deterioration and death in hospital. [2, 18] This study demonstrates that the approach is useful even in the absence of observed level of consciousness, and that physiologic data collected routinely at bedside in most facilities in Ghana and throughout Sub-Saharan Africa can generate a score (LMEWS) with high predictive value.

Serious adverse events and some portion of in-hospital mortality can be prevented by limiting human error, such as failure to recognize the early warning signs of a deteriorating patient, or failure to act on this information in a timely manner.^[19]^ MEWS is a low-cost tool that utilizes easy-to-measure bedside parameters to generate a singular value that can identify at-risk patients. This value can be used as a preset trigger in the context of a reporting algorithm.

We found that, in this setting having a LMEWS value of 4 or greater was highly associated with mortality. The area under the curve (AUC) of 0.84 for the LMEWS is consistent with good model accuracy in the discrimination of patients who are critically ill. The combination of LMEWS with clinical judgement is therefore likely to be as effective in Ghana as it has been in other similarly resourced settings.^[20]^ This is encouraging since LMEWS can be implemented without additional training of staff on how to score the level of consciousness, and without changing standardized documentation sheets already in use for patient monitoring.

The standard inpatient vital signs monitoring charts used in many Ghanaian hospitals includes a 4-hourly graphic to plot temperature, pulse rate, respiratory rate, and blood pressure. Additional parameters may also be serially recorded in some instances or centers, however the typical bedside observation chart does not record the level of consciousness for patients, as captured in the MEWS by including either the AVPU or RASS score.

Although the original description defined a “significant” MEWS as any single score ≥5, or any increase of 2+ points in patients with initial scores above 5,^[2,10]^ a cut-off of 4 was adopted for this study. Arguably, a lower threshold for detection would increase the burden of patient re-examination and re-assessment on healthcare providers, potentially making use of the score impractical in settings with severely limited human resources. The decision to adopt a cut-off score of 4 as the definition of a “significant” MEWS was based on previous work done by Gardner-Thorpe et al in 2006, which showed that raising the threshold reduces the sensitivity to unacceptable levels for patient safety, though an increase in specificity would be observed.^[16]^ Using a cut-off of 4, the proportion of individuals with a “significant” MEWS score was 33/112, and 31/112 had a “significant” LMEWS. In other words, nearly 30 percent of patients in our study would have been categorized as high-risk for clinical deterioration in the context of a MEWS-based reporting algorithm.

Interestingly, using a MEWS or LMEWS with a cut-off of ≥5 did not only yield higher discrimination, based on the C-statistics, but also had better calibration in terms of correctly assessing the risk of disease severity. Based on the receiver operating characteristics and the Hosmer-Lemeshow goodness-of-fit test, the LMEWS with a cut-off of ≥5 was superior to both MEWS and a LMEWS with a cut-off of ≥4.

Encouraging complete, accurate documentation and a standardized interpretation of vital signs with appropriate actions by nurses, doctors, and other allied staff can potentially improve the outcomes of patients admitted to hospitals, even in a setting that lacks rapid response teams. Many interventions such as fluids or antibiotics do not require advanced equipment or costly supplies, making implementation of the afferent arm of an RRS important even in settings where the efferent arm is more limited.^[20]^

This study is subject to all the limitations of a single-center, retrospective review. In addition, the study only examined vital signs collected at a single time point for each patient. Changes in serially measured physiological parameters were not evaluated. A study published by Ludikhuize et al. 2014 recommends the calculation of MEWS at least three times daily to detect the development of physiological abnormalities.^[21]^ Our study could not have detected any significant MEWS that may have developed after the first 48 hours on admission. Missing additional patients who may have worsened later and then died would bias the study towards the null hypothesis. This makes our study design a conservative one, with results consistent with other published literature on the topic.^[2, 16]^

More prospective research is needed to help define its utility and impact on patient outcomes and patient care; this study suggests that such future efforts should also consider evaluation of LMEWS because of its simplicity.

## CONCLUSION

This study is the first to examine the ability of an early warning system to predict inpatient mortality based on routinely collected clinical data in a low-resource setting. Early recognition of clinical status decline is critical even in low-resource settings, where bedside interventions may prevent ICU admissions and disease complications including death. Though the MEWS system provides good discrimination, the LMEWS provides better discrimination and calibration in the prediction of mortality and can identify critical illness among inpatients with primarily medical diagnoses. Additional prospective studies will be useful to validate LMEWS among other categories of in-patients, and to investigate its impact on health resource allocation and clinical outcomes in low-resource settings.

**What is already known on this topic;**

Changes in physiological parameters can be used to predict serious adverse events (SAEs).

MEWS is a validated composite score that identifies in-patients at risk of deterioration.

**What this study adds;**

Validation of an early warning score using currently collected physiological parameters (LMEWS).

Validation of the traditional MEWS in a resource-limited environment

## Data Availability

Data is available upon request

## CONSENT FOR PUBLICATION

Yes, consent is granted for publication.

## AUTHORS’ CONTRIBUTION

EJA was responsible for the concept, study design, part data collection, analysis and interpretation, manuscript writing. PA, JSM, MC & SS participated in the concept, critical revision and review of the manuscript. PA, JSM & MC doubled as academic mentors, while SS doubled as on-site preceptor.

## ACKNOWLEDGEMENTS

We are grateful to the KBTH, medical department staff and clientele for making patient charts available for data collection. Drs. Benjamin Sena Fenu & Oforiwaa Amoah who contributed to the data collection and on-site follow-up for ethical and technical committee approval. Mr. Nortey who was instrumental in guiding us through the ethical review processes for the KBTH. George Mwinnyaa & Seth Bennett reviewed the statistical analysis.

## LIST OF TABLES, GRAPHS, AND FIGURES

**GRAPH 1:**
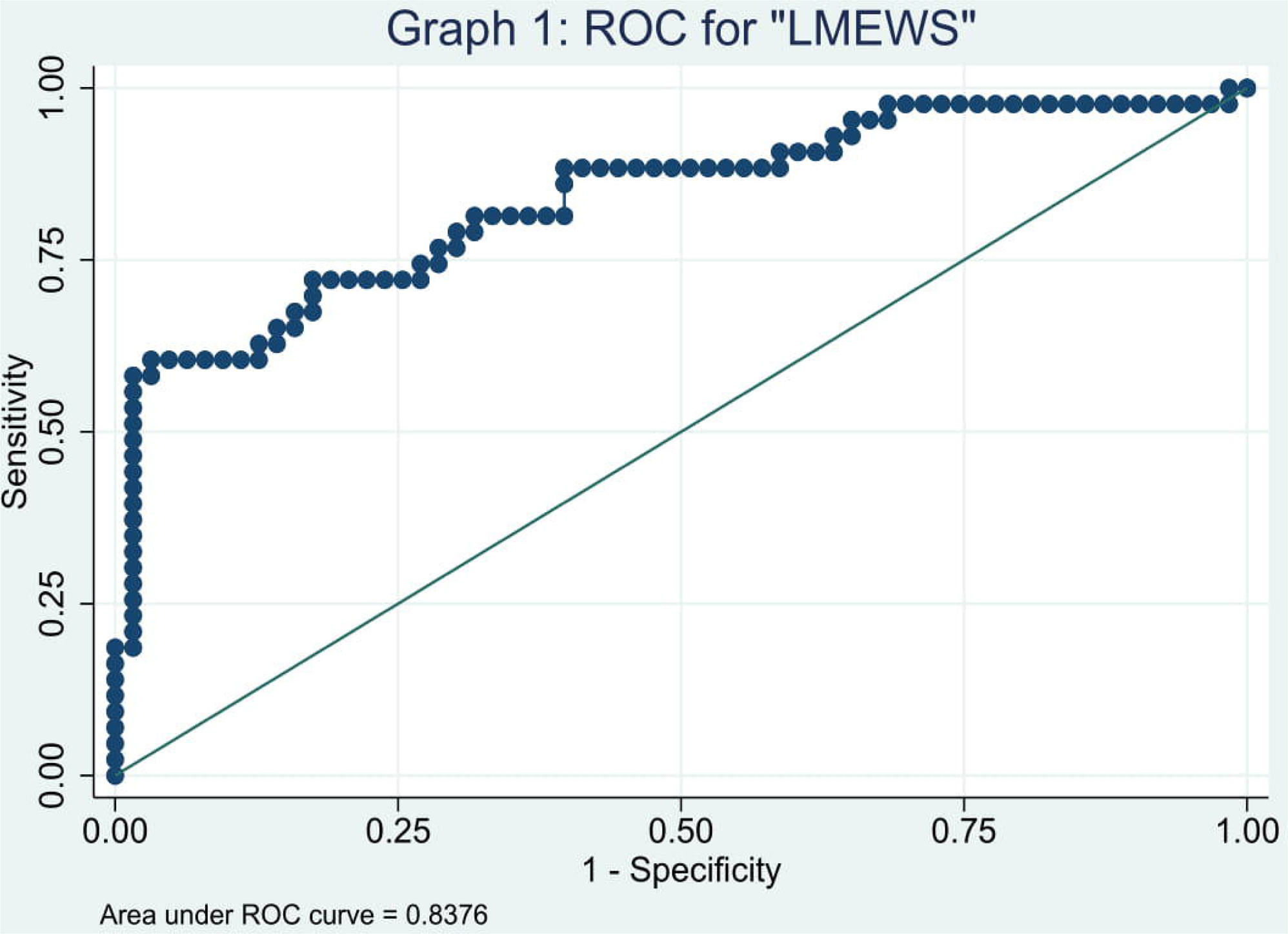
Receiver Operator Characteristic (ROC) for Limited MEWS using a cut-off of 4

**GRAPH 2:**
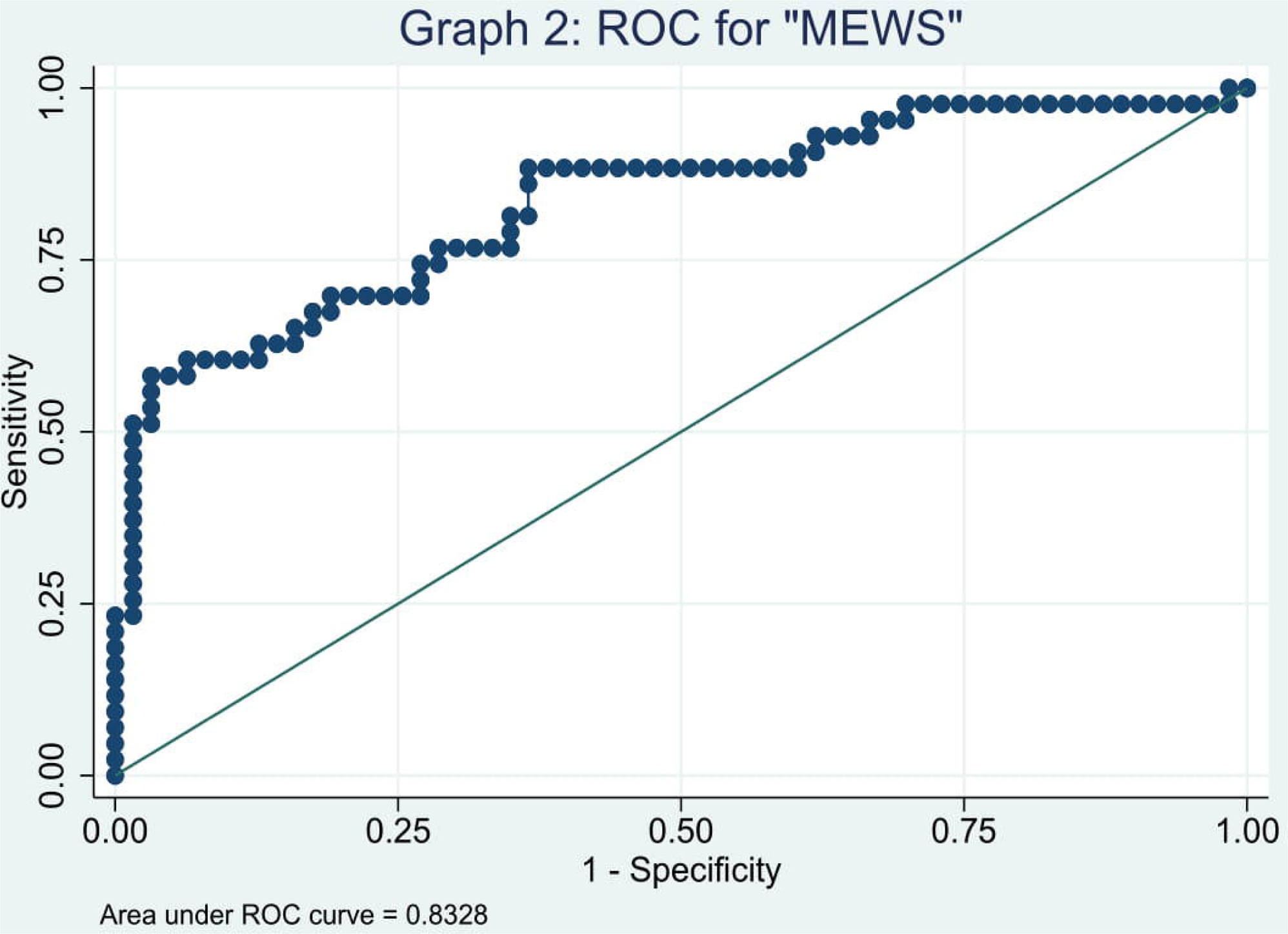
Receiver Operator Characteristic (ROC) for Full MEWS using a cut-off of 4

**GRAPH 3:**
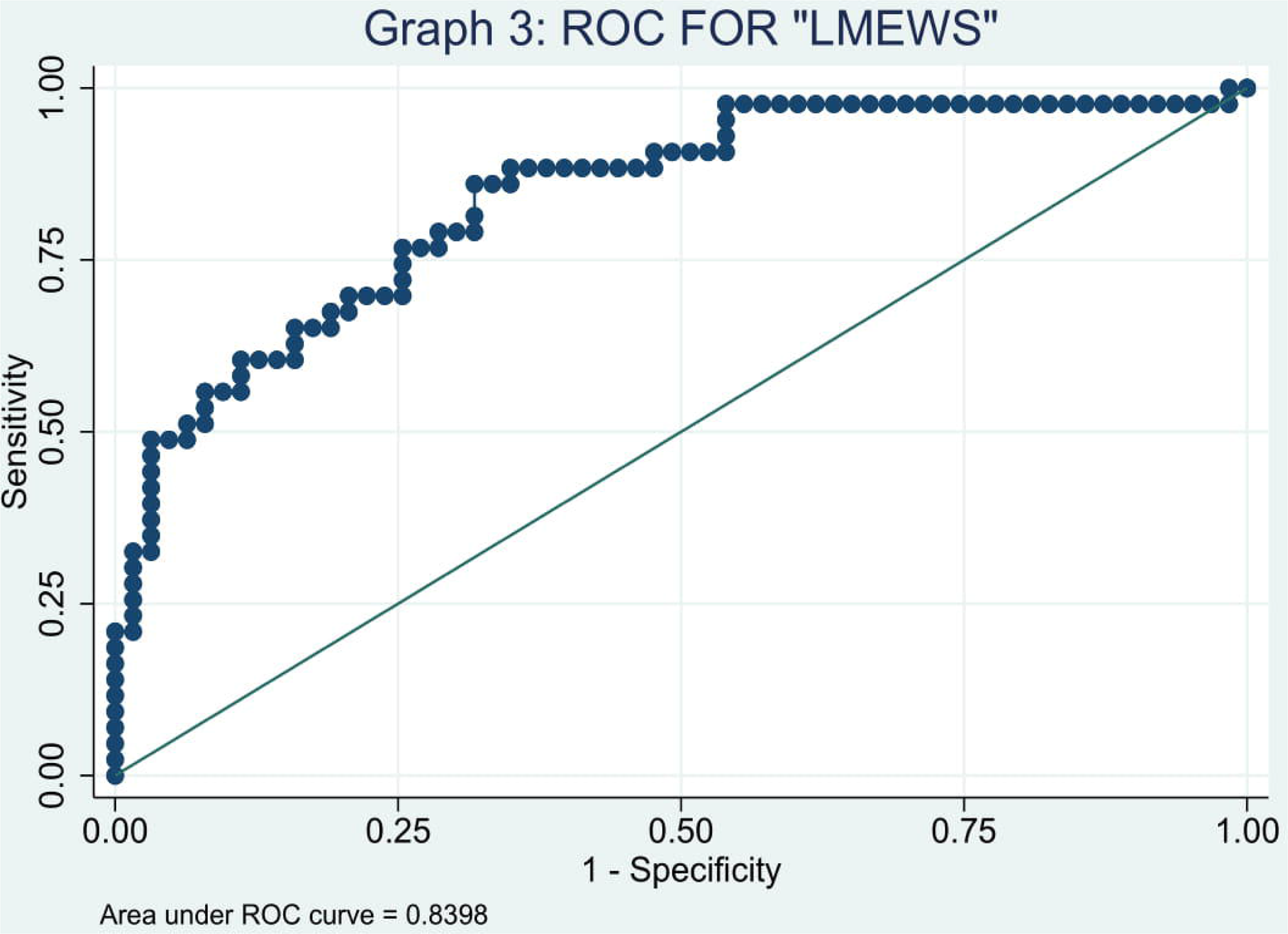
Receiver Operator Characteristic (ROC) for Limited MEWS using a cut-off of 5

**GRAPH 4:**
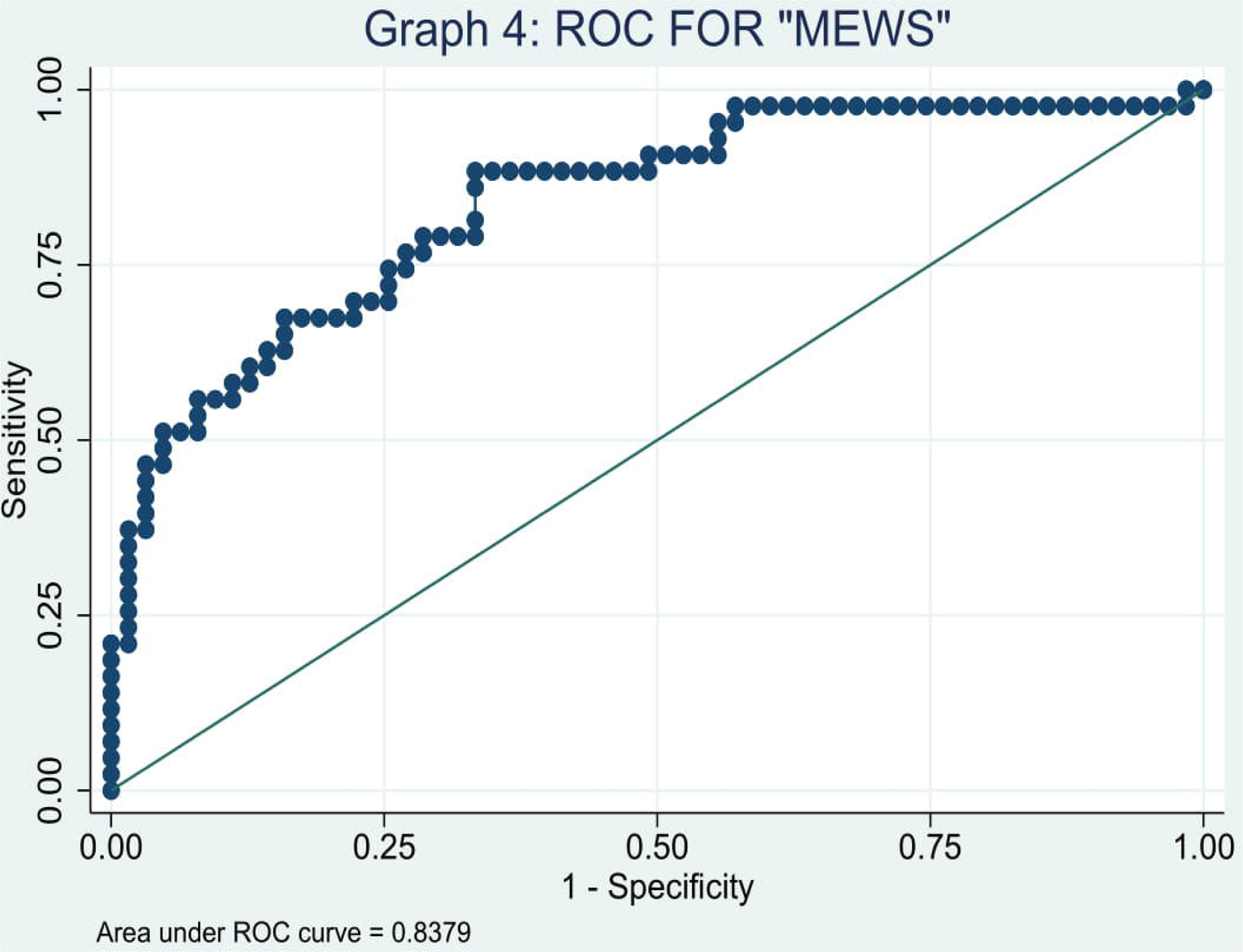
Receiver Operator characteristic (ROC) for Full MEWS using a cut-off of 5

